# 10.4 Million Children Affected by COVID-19-associated Orphanhood and Caregiver Death: An Imperative for Action

**DOI:** 10.1101/2022.05.08.22274788

**Authors:** Susan Hillis, Joel-Pascal Ntwali N’konzi, William Msemburi, Lucie Cluver, Andrés Villaveces, Seth Flaxman, H. Juliette T. Unwin

**Affiliations:** Global Reference Group on Children Affected by COVID-19, University of Oxford, UK; African Institute for Mathematical Sciences, Rwanda; World Health Organization Division of Data, Analytics, and Delivery for Impact; Department of Social Policy and Intervention, Oxford and Department of Psychiatry and Mental Health, University of Cape Town; CDC COVID-19 Response Team, U.S. Centers for Disease Control and Prevention, Atlanta, Georgia; Department of Computer Science, University of Oxford, Oxford, UK; MRC Centre for Global Infectious Disease Analysis and the Abdul Latif Jameel Institute for Disease and Emergency Analytics (J-IDEA), School of Public Health, Imperial College London

## Abstract

The new WHO estimates for COVID-19 excess deaths allow us to generate updated and more accurate models of COVID-19 associated orphanhood and caregiver loss. Using methodology established in prior studies, we combine age-specific fertility and excess death estimates from January 2020 to May 2022. We find 10.4 million children have lost a parent or caregiver due to COVID-associated excess deaths, and 7.5 million children have experienced COVID-associated orphanhood. Without supportive intervention, caregiver loss can bring severe risks of poverty, school dropout, sexual exploitation, and mental health distress. It is essential that evidence-based care for these children is integrated into all national response plans as a caring action to protect children from immediate and long-term harms of COVID-19.

## Introduction

Emerging global modeling data on COVID-19-associated excess deaths suggests the pandemic mortality toll is more than twice the number of reported COVID-19 deaths.^1-3^ For the first time, the availability of these comprehensive data enables us to update global minimum estimates of pandemic orphanhood and caregiver death among children.^4,5^ Parent or caregiver death is permanent; consequences for children can be devastating and enduring, including institutionalization, abuse, traumatic grief, mental health problems, adolescent pregnancy, poor educational outcomes, and chronic and infectious diseases.^4,5^ Global totals and country comparisons were previously hampered by inconsistencies in COVID-19 testing and incomplete death reporting. The new orphanhood estimates we derive here based on excess mortality provide a comprehensive measure of the pandemic’s long-term impact on orphanhood and caregiver loss.

## Methods

Using previous methodology for combining age-specific death and fertility rates,^4,5^ we update our estimates of parent and caregiver loss to present excess mortality-derived estimates for every reporting country, using data from the World Health Organization (WHO), The Economist, and The Institute of Health Metrics and Evaluation (IHME).^1-3,6^ We replace COVID-19 deaths in previous logistic models with the maximum of excess deaths and COVID-19 deaths, for two periods: January 1, 2020, through December 31, 2021, and January 1, 2020, through May 1, 2022 (Supplementary Methods (SM)). We use bootstrapping to calculate uncertainty around our estimates from both fertility and death data (SM). We show 95% credible intervals in brackets. We present WHO regional and national estimates; trends were similar across datasets.

## Results

Using WHO excess mortality (more conservative than IHME and Economist findings), we estimate that 10,400,000 children lost parents or caregivers (Table 1), and 7,500,000 children experienced pandemic-associated orphanhood, through May 1, 2022. These children are not equally distributed, with greater numbers affected by orphanhood and caregiver loss in the South-East Asia (40.9% [35.6%-46.5%] and Africa (23.7% [18.6%-27.2%] WHO regions compared to the Americas (14.1% [12.7%-15.9%]), Eastern Mediterranean (14.7% [13.0%-16.3%] European (4.8% [4.5%-5.3%]) and Western Pacific regions (1.8% [1.8%-1.9%]) through May 1, 2022 (Figure 1A). Similarly, variation in estimates arises at national levels, with India (3,490,000), Indonesia (660,000), Egypt (450,000), Nigeria (430,000), and Pakistan (410,000) worst affected, through May 1, 2022 (Figure 1B). Among the WHO regions most affected, countries with the highest numbers of bereaved children in South-east Asia include India, Indonesia, Bangladesh, Myanmar, and Nepal; and in Africa, include Nigeria, Democratic Republic of Congo, Ethiopia, South Africa, and Mozambique. Our updated Orphanhood Calculator^6^ provides these new numbers by country.

**Table 1:**
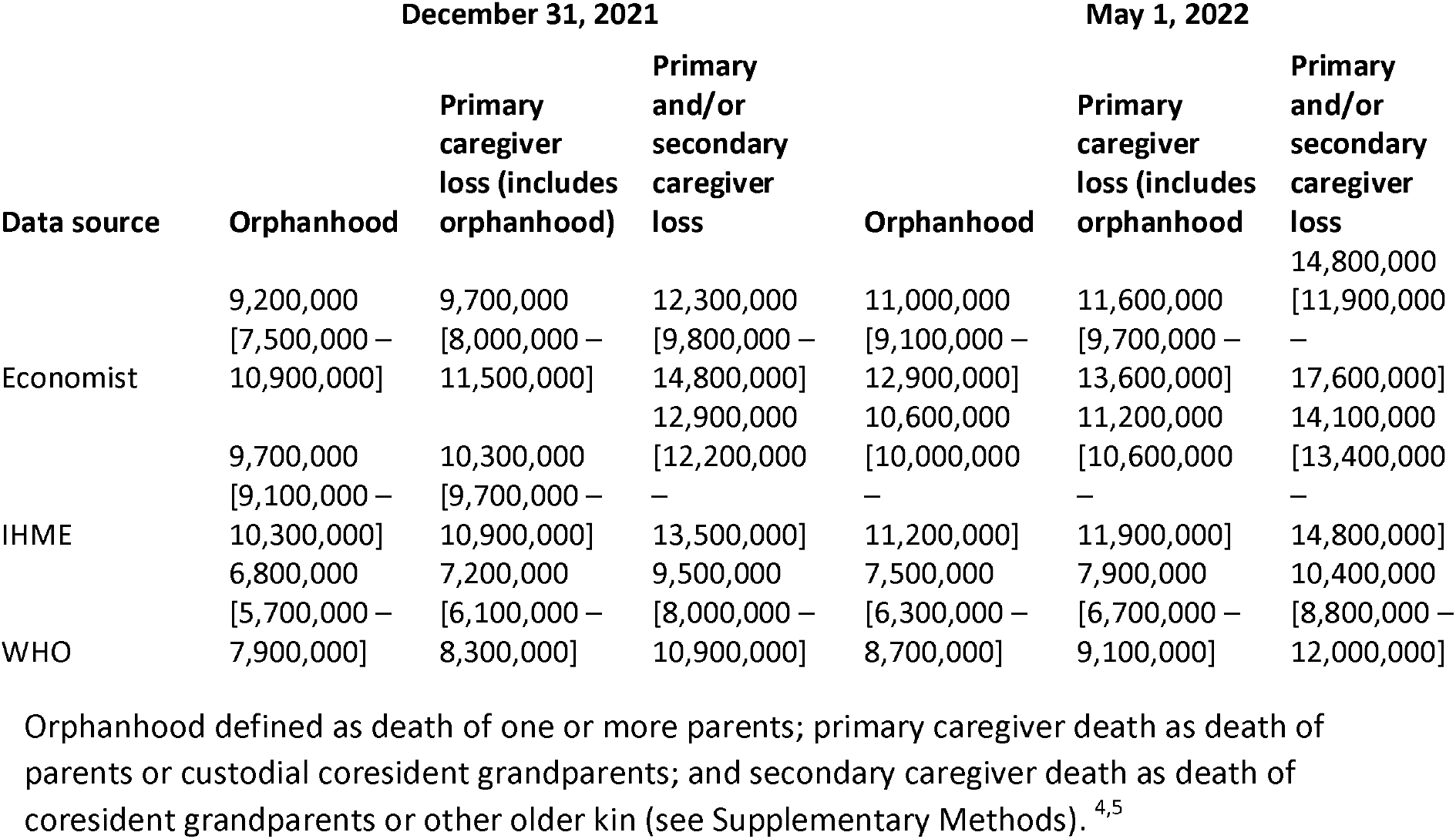
Estimates of orphanhood and caregiver loss using excess death estimates from The Economist, IHME and WHO through December 31, 2021 (end of reporting period for IHME and WHO datasets) and adjusted using Johns Hopkins University data through May 1, 2022. ^5 6^

**Figure 1:**
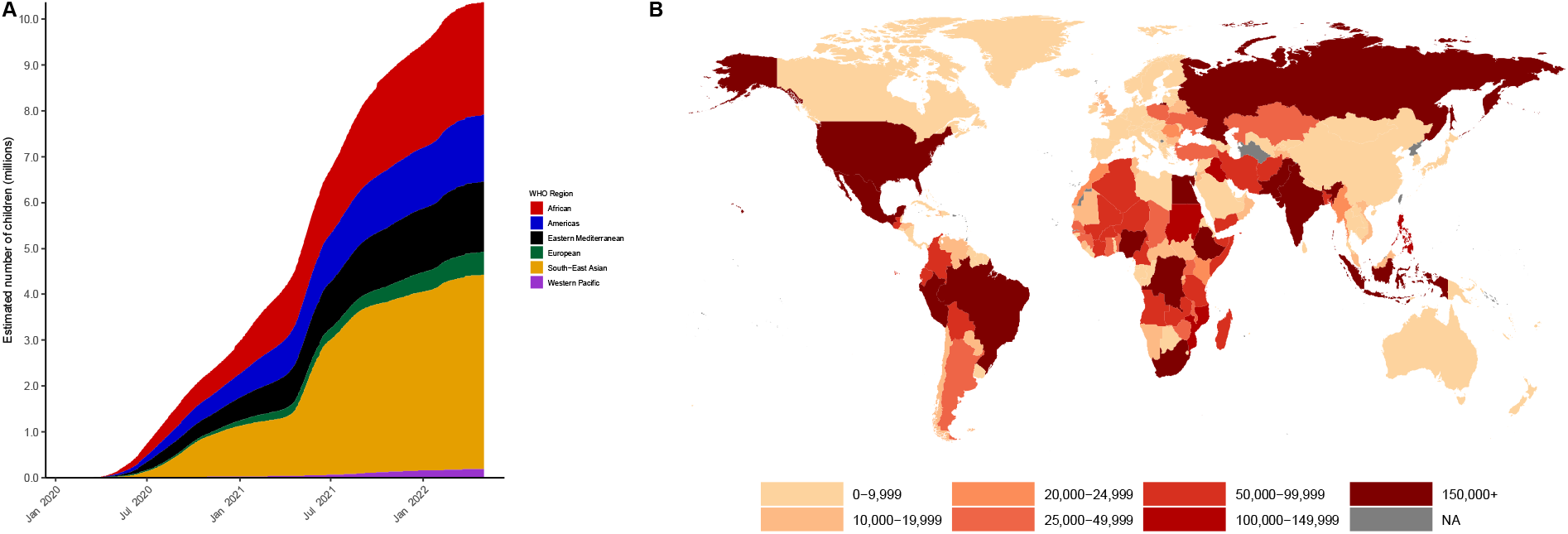
WHO Regional (A) and national (B) estimates of orphanhood, primary and/or secondary caregiver loss through May 1, 2022, using WHO methods for estimates of excess death adjusted using Johns Hopkins University data. (Classification of countries by WHO region as previously reported.^5^) Countries with negative estimates occur in countries with negative excess deaths – therefore fewer children have lost parents or caregivers during this time than expected.

## Discussion

The global pandemic of COVID-associated orphanhood and caregiver death has left an estimated 10.4 million children bereaved of their parents and caregivers. These new data on the pandemic’s consequences for children align with ‘Saving Lives Now,’ a key priority for the upcoming Presidential Global COVID-19 Summit. While billions of dollars are invested in preventing COVID-19-associated deaths, little is being done to care for children left behind. However, reports of billions of dollars invested in supporting AIDS-orphaned children showcase successful solutions ready for replication.^5^ Only two countries: the US^1^ and Peru^2^ – have made evidence-based national commitments to address COVID-associated orphanhood. Urgently needed pandemic responses can combine equitable vaccination with life-changing programs for bereaved children. Given the magnitude and life-long consequences of orphanhood, integration into every national pandemic response plan of timely care for these children will help mitigate the life-long consequences. Evidence highlights three essential components: 1) prevent death of caregivers by accelerating vaccines, containment, and treatment; 2) prepare families to provide safe and nurturing alternative care; and 3) protect orphaned children through economic support, violence prevention, parenting support and ensuring school access. Effective, caring action to protect children from immediate and long-term harms of COVID-19 is an investment in the future and a public health imperative. ^5^

## Data Availability

All data produced are available online at https://github.com/ImperialCollegeLondon/covid19_orphans

## Data Availability

All data produced are available online at https://github.com/ImperialCollegeLondon/covid19_orphans

## Acknowledgments

We acknowledge Jon Wakefield for his help with accessing data and comments on the manuscript.

## Supplementary Methods

We adapt previously published methods^4,5^ to estimate orphanhood and caregiver loss from excess death estimates made by The Economist^2^, The Institute of Health Metrics and Evaluation (IHME)^1^, and the World Health Organization (WHO).^3^ We consider three different categories: orphanhood, primary caregiver loss and primary and/or secondary caregiver loss. We defined orphanhood using UNICEFs definition of the loss of one or more parents; primary caregiver death as death of parents or custodial coresident grandparents (providing care for children in the absence of parents); and secondary caregiver death as death of coresident grandparents or other older kin (providing care through involvement or resources). ^4,5^

In Unwin et al.,^5^ we use a logistic regression model to calculate orphanhood and caregiver loss ratios to deaths based on total fertility rate. We fit this model to data calculated from 21 study countries (Argentina, Brazil, Colombia, England & Wales, France, Germany, Kenya, Malawi, Mexico, Nigeria, India, Iran (Islamic Republic of), Italy, Peru, the Philippines, Poland, Russian Federation, Spain, South Africa, United States of America, and Zimbabwe). In the current report, instead of using COVID-19 death data from Johns Hopkins University for all countries as in our most recent report,^5^ we multiply our orphanhood and caregiver loss to death ratio by the maximum between excess deaths and COVID-19 deaths for every country with data, from two time periods: January 1, 2020, through December 31, 2021 (end of reporting period from IHME^1^ and WHO^3^ datasets) and January 1, 2020, through May 1, 2022. This enables us to account for underreporting in COVID-19 data and is consistent with previous methodology.^4,5^

We use a death adjustment factor for each country to update our data outside the time-period for the IHME and WHO data, calculated on December 31, 2021, when we have data for both excess and COVID-19 deaths. Specifically, we assume the ratio of excess deaths to COVID-19 deaths through Dec 31, 2021, was consistent with the ratio between January 1, 2022 and May 1, 2022, for WHO and IHME estimates. We, therefore, multiplied the new COVID-19 deaths by the ratio for the prior period to generate an estimate of country-specific excess deaths for this new period. If COVID-19 deaths were greater than excess deaths, we used the reported value of COVID-19 deaths. The Economist data are released weekly and available through May 1, 2022, so no data adjustment factor is needed. The Economist dataset includes 224 countries, IHME includes 190 countries, and estimates based on WHO methods include 181 countries, due to how the data are grouped.

We consider uncertainty in both the total fertility rate and excess deaths using bootstrapping, similar to methods previously described^4,5^. For all three data sets, we assume the total fertility rate is normally distributed with the standard deviation based on the lower and upper bounds. We do not allow the total fertility rate to vary for our 21 study countries and use the ratios generated for the study opposed to ones estimated from the logistic model. For the IHME and Economist data we also assume the number of excess deaths were normally distributed with the standard deviation based on the lower and upper bounds. However, for the WHO data we sample the excess deaths with replacement from their 1000 Markov Chain Monte Carlo draws. Constant with previous methodology^4^, we assume deaths were constant across all samples if COVD-19 deaths were greater then excess death. We use 5,000 samples for our bootstrap since at this number of samples, the error has begun to converge.^3^

## Footnotes

1 Memorandum on Addressing the Long-Term Effects of COVID-□19. Accessed on April 21, 2022. Available at: https://www.whitehouse.gov/briefing-room/presidential-actions/2022/04/05/memorandum-on-addressing-the-long-term-effects-of-covid-19/

2 Decreto Supremo que aprueba las Disposiciones Complementarias para el otorgamiento de la Asistencia Económica a favor de las niñas, niños y adolescentes cuya madre o padre o ambos hayan fallecido durante el periodo de Emergencia Sanitaria por causa de la COVID-19 decreto supremo N° 002-2021-mImp. Accessed on May 3, 2022. Available at: https://cdn.www.gob.pe/uploads/document/file/1718741/ds-002-2021-mimp.pdf

3 Christopher J. Freitas, The issue of numerical uncertainty. Applied Mathematical Modelling. Volume 26, Issue 2, 2002, Pages 237-248, ISSN 0307-904X, https://doi.org/10.1016/S0307-904X(01)00058-0.

